# Feasibility of Social Media Recruitment for Orofacial Cleft Genetic Research

**DOI:** 10.1101/2021.03.09.21253219

**Authors:** Grace Carlock, Kelly Manning, Elizabeth J. Leslie

## Abstract

**Objective:** This study assessed the feasibility of unpaid social media (SM) advertising to recruit participants affected with an orofacial cleft (OFC) for a genetic study.

**Design:** This paper is a retrospective analysis of study recruitment based on enrollment and participation in a genetic study. Participants completed a series of enrollment surveys, provided saliva samples for genetic analysis, and completed post-participation feedback surveys.

**Participants:** Participants signed up for the study following SM advertisement. Participants were eligible if: they or a minor in their care were affected by an OFC, the affected participant was not adopted, and the mother of the affected individual had not taken anti-seizure medication during pregnancy. 313 individuals completed initial screening surveys; 306 participants were eligible. 263 individuals completed all online surveys and were sent DNA sample kits. 162 subject DNA samples were returned.

**Main Outcome Measures:** Success of recruitment was evaluated as number of enrolled participants and return rate for DNA samples.

**Results:** 263 OFC-affected individuals completed the screening process in the first 12 months of recruitment. 162 of 263 affected subject DNA samples were returned within 12 months of sending, for a return rate of 62%. Approximately two-thirds (66.3%) of all returned samples were sent back within the first 6 weeks after receiving DNA kits.

**Conclusions:** Unpaid SM advertising enabled the recruitment of a large cohort of participants in a short time (12 months). SM recruitment was inexpensive and effective for recruiting participants for a genetic study.

## Introduction

Orofacial clefts (OFCs) are one of the most common congenital anomalies, occurring in approximately 1 in 1000 live births.^1^ OFCs typically present as cleft lip, cleft palate, or cleft lip and palate. OFCs can present as isolated congenital anomalies (non-syndromic) or, when in conjunction with other structural or cognitive anomalies, as part of a syndrome. Non-syndromic OFCs represent roughly 70% of cleft cases.^2^ OFCs represent a major source of morbidity in affected children and is a source of significant financial and social burden for families.^1-3^ Affected individuals and their caregivers can experience lifelong social stigma and mental health challenges, resulting in the need for robust social support.^1-4^

The popularity of social media (SM) sites exploded in the years since the introduction of Facebook and MySpace in the early 2000s. Facebook is now the highest traffic SM site in the world, with over 2.7 billion active monthly users.^5^ Individuals and families affected by medical issues are increasingly turning to social media for support and camaraderie. Studies on the use of SM by patients with rare diseases show that they seek out online groups for information, social support, and advice.^4,6-11^ A 2020 study by Çinar et al found patients and families affected by OFCs share similar motivations for using SM support groups.^4^ Two studies in 2018 identified learning about new research as an impetus for participation in OFC SM groups.^9,10^

Because social media users are geographically and demographically diverse, they also represent a convenient source of potential research participants. Rare disease researchers were among the first to successfully tap into SM. In 2014, Schumacher et al utilized SM to assemble the largest-ever cohort of individuals with Fontan-associated protein losing enteropathy and plastic bronchitis.^12^ Multiple studies of auto-immune hepatitis mobilized Facebook and other SM to successfully enroll participants who were geographically distant from primary study centers.^11,13,14^ SM was also used to facilitate recruitment of individuals who have not yet developed cancer (previvors), an oft-missed demographic in the study of Lynch syndrome.^15^

Recruitment via SM offers a number of advantages. It is faster and, in most cases, less expensive than traditional study advertising methods.^11-21^ Participants are not limited by geographic proximity to study centers. Traditional barriers, such as conflicting work and study center schedules or travel time, are not issues with SM-based recruitment, which allows patients to complete study surveys or sample collections on their own time in the privacy of their home.^6,11-21^ In addition, studies that recruit via Facebook and other SM sites are consistently geographically diverse and typically demographically representative of the larger patient population.^17,21^ Those studies that are not demographically representative tend to be overly white and female.^17,21^

While many studies use paid advertisements on SM, only a few studies have examined the efficacy of unpaid study advertisements in SM support groups, especially when collecting physical samples.^13,16,17,20,21^ We established the Emory Cleft Project (ECP) at Emory University to study the genetic causes of OFCs, which requires participants and their families to fill out a series of background surveys and submit saliva samples for genetic analysis. The ECP primarily utilizes social media (SM) advertising to recruit participants to the study, with some limited clinic-based recruitment. Here, we present the results of the ECP’s study recruitment using unpaid advertising in Facebook support groups for individuals and families affected by OFCs. This paper evaluates the feasibility of unpaid SM advertisement as a recruitment tool for genetic studies.

## Methods

### Recruitment

Prior to conducting study procedures, IRB approval was obtained through the Emory University Institutional Review Board (IRB#00105750). Recruitment for the study involved identifying active Facebook support groups for OFCs using the Facebook search function. Several support groups were identified for initial outreach based on number of members and activity level as measured by number of daily posts - groups with average post activity of at least one post per week were considered “active” groups. Direct messages were sent to the administrators of relevant active groups with a request to post on their support group pages. Post content included a study flier containing contact information, a brief summary of the study, and a link to complete a brief online screening survey. A logo was created for the Emory Cleft Project and included on study fliers, posts, and other content to generate consistency and professionalism and to improve recognition.

Interested individuals used the provided link to complete a short screening survey to indicate interest and to be evaluated for eligibility. They could also contact the study staff directly for further information or to be screened by phone or email. Participant data collection, storage, and survey distribution was managed using REDCap electronic data capture tools hosted at Emory University.^22,23^ REDCap is a Health Insurance Portability and Accountability Act of 1996 (HIPAA) compliant browser-based project management platform designed to support data capture for research studies, including building and managing databases and surveys online.

### Eligibility and Enrollment

In order to be eligible to participate, individuals were required to have a congenital orofacial defect of the lip and/or palate, including overt and microforms (e.g., submucosal cleft palate or microform cleft lip). Individuals without an OFC but with congenital lower lip pits and a diagnosis of Van der Woude Syndrome were also eligible. Additionally, it was necessary that the individual and/or his/her parent or legal guardian be able to read and write in fluent English and have access to family medical history for at least one biological parent. Individuals with certain syndromes or medical conditions in addition to the primary craniofacial defect were excluded from the study.

Screening was conducted primarily through the completion of a brief online screening survey of the inclusion and exclusion criteria by the interested individual, which would then be reviewed for eligibility. Occasionally, the screening would be conducted by a study staff member over the phone or email using a prepared screening script. Once an individual was determined to be eligible, a unique study ID was assigned, and the participant received an email with instructions and a link to complete the online consent form and begin the online surveys. All participants had to complete written informed consent forms in an online format before taking part in the study.

### Study Procedures

After enrollment in the study and completion of the online consent form, participants completed four online surveys. The first assessed basic demographic characteristics, other medical conditions, and details about the subject’s OFC diagnosis. The second survey captured family information to construct a pedigree as well as history of craniofacial anomalies within the family. The third survey captured information about personal and family medical history, including other anomalies associated with orofacial clefts. The final survey asked about potential environmental exposures and behavioral risk factors during pregnancy, including tobacco and alcohol use. Participants who did not complete their surveys within one week received an automated reminder email, up to 3 times. Completed surveys were reviewed by study staff for completeness and accuracy, and compensation was provided in a follow-up email to the participant. Compensation for survey completion was $25 USD in the form of an Amazon gift card.

After survey completion, sample collection kits were sent via FedEx to the participant’s residence. Samples were collected from the participant, as well as both biological parents when possible. Occasionally, based on medical history or other factors, additional family members invited to participate and to provide a sample. The sample collection kit contents included: Oragene saliva kits for each eligible individual, instructions for collection, consent forms for all additional participants who had not consented online, return shipment packing materials and return postage. Once samples were returned and checked in, participants were contacted to confirm study completion and final compensation of $25 USD per sample provided was distributed. Reminders for unreturned shipments were sent via email approximately 1 month after shipment, with follow-up reminders every 4-6 weeks for about 6 months, after which point if there was no further response from the participant they were assumed to be lost-to-follow-up.

### Post-Participation Feedback Survey

A feedback survey about study participant experiences was sent to families who had completed the online surveys and sample collection for the study at the time the survey was distributed. Participation in this survey was voluntary and was not compensated. The purpose of the survey was to gather information about participant experiences, including ease of participation, motivation for participating, feedback about the study process, and preferences for dissemination. The survey also asked about previous experiences with research participation, genetic testing, and genetic counseling.

## Results

Recruitment for the Emory Cleft Project is ongoing. For the purposes of this analysis, only participants who were screened and/or enrolled from the start of recruitment in March 2019 through the end of 2019 were included. This date cutoff for inclusion in analysis was based on expected completion time for sample return after enrollment, allowing for a minimum of 12 months from initial consent to return the samples.

### Enrollment and Participation

A total of 313 individuals from 267 families completed the online screening process, of which 306 (98%) individuals from 260 families were determined by the study staff to be eligible for the study and were invited to participate. Of the eligible individuals, 269 study subjects from 226 families completed the online consent process (88%) - 263 completed the online surveys (98%), while 6 consented individuals initiated but did not complete the surveys. Most of the consents and corresponding surveys (219; 81.4%) were completed by the parent/guardian of the enrolled minor study subject. The remaining consents and surveys (50, 18.6%) were completed by adult study subjects.

DNA sample kits were shipped to the 263 study subjects from 220 families who completed the surveys. In addition to DNA sample kits for study subjects, sample kits were sent to biological parents and other affected family members, when applicable. Returned samples came from 133 families. Family structures included 5 singleton cases, 11 maternal dyads, 94 complete trios (proband and both parents), and 21 nuclear or multiplex families. An overview of the study participation and drop-off at each stage of the study process is summarized below in Figure 1.

**Figure 1.**
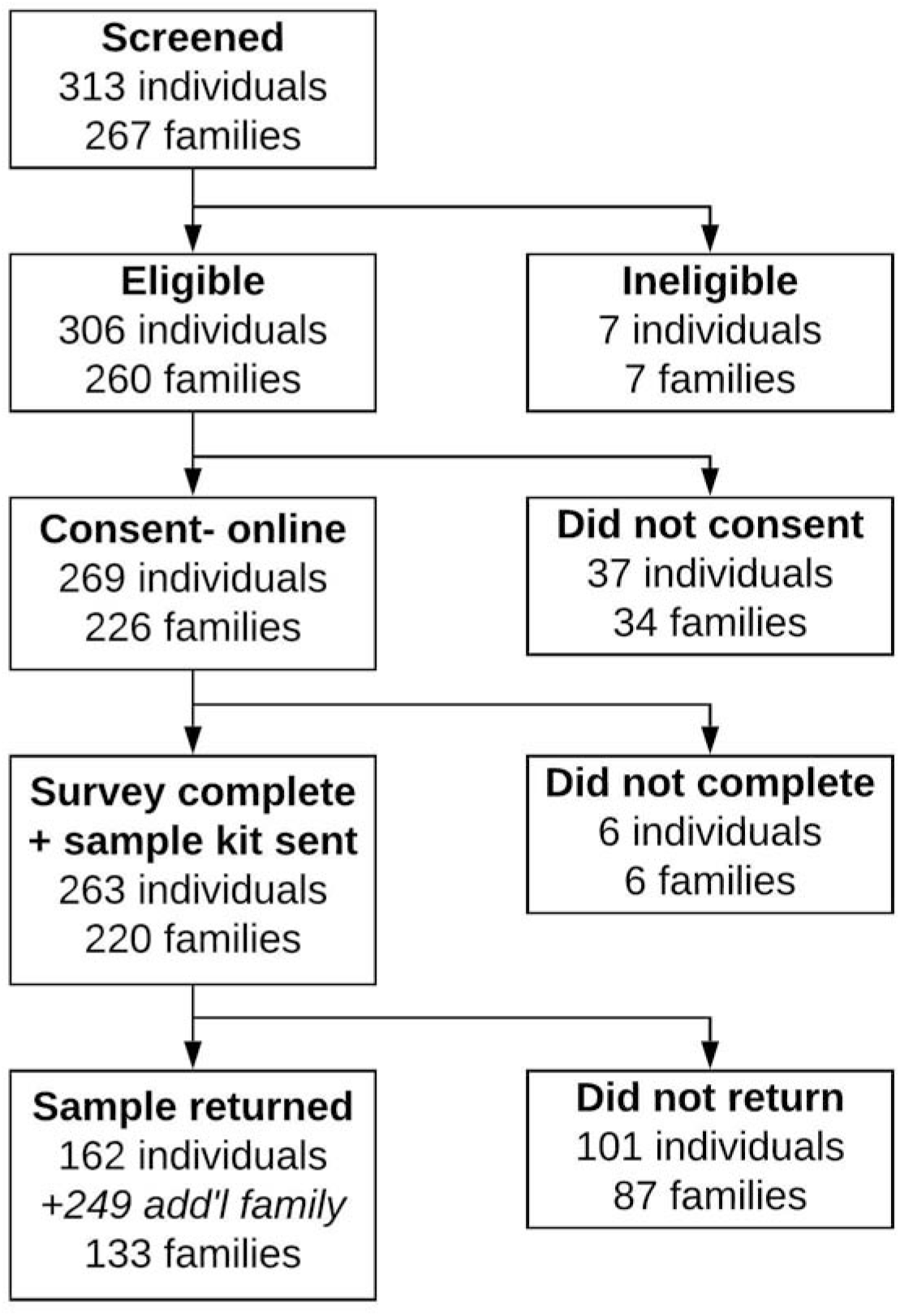
Study Participation and Dropoff from March 2019 to December 2019

### Sample Return

Samples were received from a total of 411 participants, including 196 male samples (47.7%) and 215 female samples (52.3%). Of the kits that were returned (which accounted for only 60% of shipped kits), 25% were returned within 2 weeks of shipment, and 45% were returned within one month. Median return time for sample kits was 34 days, or approximately 4-5 weeks. Returns tapered over time – by 3 months (12 weeks) from the shipment date, 80% of the returned samples had been received, and only 8% of returned samples arrived after 6 months (Figure 2). It is worth noting that 16 samples were received more than a year after they were shipped, with the longest return time of 74 weeks (17 months) from shipment date. As of the writing of this manuscript, approximately 13 months have passed since the last shipment of sample kits in 2019, so there may still yet be returned samples that will not be counted here, although we do not expect there to be a substantial number.

**Figure 2.**
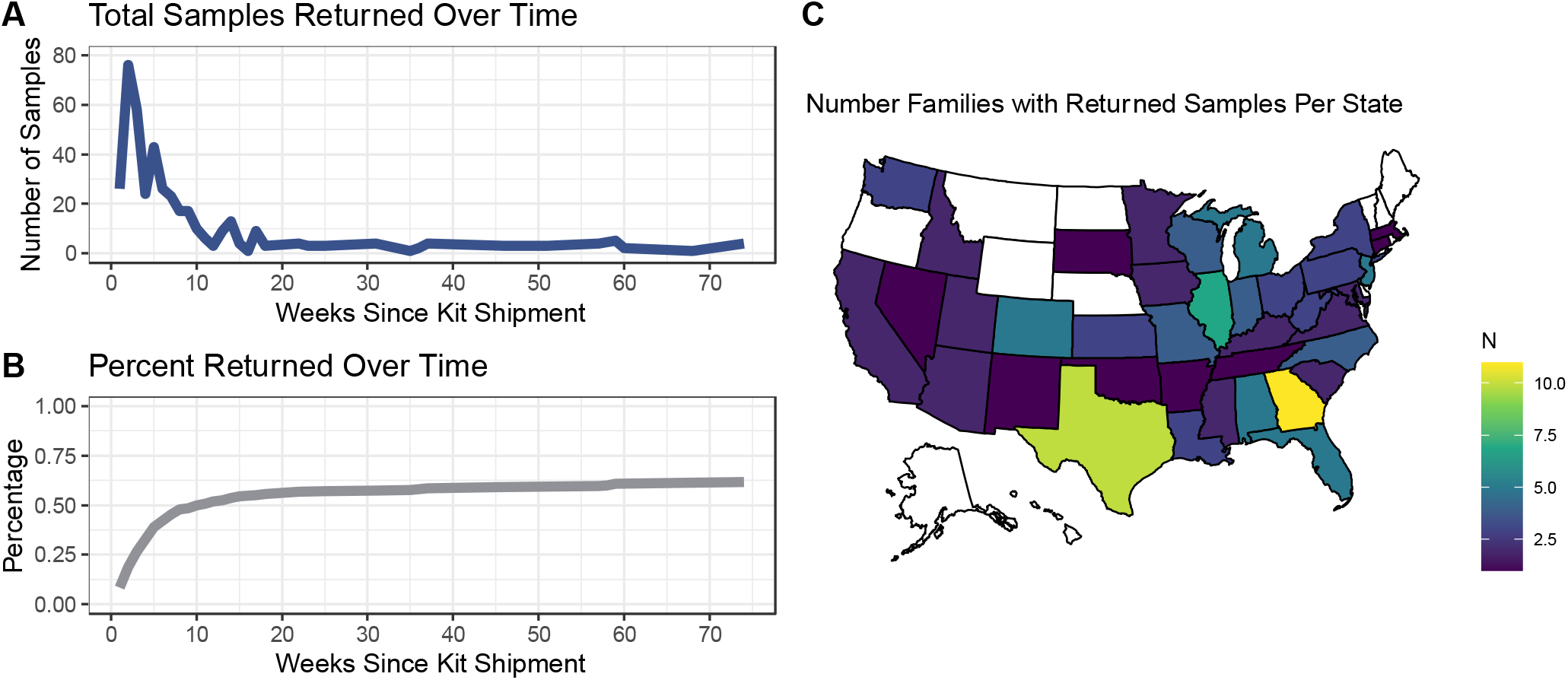
Sample return rate and geographic distribution of participation in the United States. (A) Number of samples returned per week after sample kit shipment. (B) Cumulative percentage of shipped sample kits returned over time. (C) Geographic distribution of families returning samples in the United States. States colored white have no enrolled participants during 2019.

### Geographic Distribution

A total of 226 families consented into the study and provided geographic information (state or country). The majority (205 families) were located in the United States across 44 states. Twenty-one international families participated from 7 countries (Australia, Canada, France, Germany, Ireland, the Philippines, and New Zealand). Samples were received from 133 total families, 120 of which resided in the United States across 38 states (Figure 2). Twelve international families who returned samples from 6 countries (Australia, Canada, France, Ireland, New Zealand, and the Philippines). Although recruitment was conducted remotely, the state where the study was based (Georgia) had the highest enrollment, with 17 families consented and 11 families with returned samples.

### Demographic Characteristics

A total of 518 study subjects were consented into the study in 2019. Six did not complete the surveys or provide samples; 101 completed surveys but did not return samples, 162 completed surveys and returned samples, and 249 were family members who consented and provided samples only. Summary statistics of study subjects are presented in Table 1. Of the 411 study subjects who provided samples, 158 subjects from 130 families were reported to have an orofacial cleft (OFC). Two families were included in the study where the proband did not have an OFC but had lower lip pits, indicative of Van der Woude Syndrome.

**Table 1:** Summary Statistics.

| **Table 1: Summary Statistics** |  |  |  |
| --- | --- | --- | --- |
|  | **All with Samples** | **OFC Samples** | **Enrollees** |
| **Total** | 411 | 158 | 240 |
| **Sex (n, %)** |  |  |  |
| Male | 196 (47.7%) | 75 (47.5%) | 14 (5.8%) |
| Female | 215 (52.3%) | 83 (52.5%) | 226 (94.2%) |
| **Age (mean, SD)** | 11.9 (15.5%) | 11.7 (15.0) | 35.4 (8.0) |
| **Ethnicity (n, %)** |  |  |  |
| Hispanic or Latino | 26 (6.3%) | 12 (7.6%) | 14 (5.8%) |
| Not Hispanic or Latino | 385 (93.7%) | 146 (92.4%) | 226 (94.2%) |
| Unknown | - | - |  |
| **Race (n, %)** |  |  |  |
| American Indian or Alaska Native | 3 (0.7%) | 3 (1.9%) | 4 (1.7%) |
| Asian | 7 (1.7%) | 2 (1.3%) | 4 (1.7%) |
| Black or African American | 10 (2.4%) | 4 (2.5%) | 6 (2.5%) |
| More than one race | 8 (1.9%) | 3 (1.9%) | 4 (1.7%) |
| Prefer not to answer | 1 (0.2%) | - | - |
| White | 382 (92.9%) | 146 (92.4%) | 221 (92.1%) |
| Unknown | - | - | 1 (0.4%) |
| **Cleft (n, %)** |  |  |  |
| Yes | 158 (38.4%) |  | 42 (17.5%) |
| No | 253 (61.6%) |  | 98 (82.5%) |
| **Cleft Type (n, %)** |  |  |  |
| Total | 158 | 158 | 42 |
| CLP | 104 (65.8%) | 104 (65.8%) | 26 (61.9%) |
| CL | 29 (18.4%) | 29 (18.4%) | 11 (26.2%) |
| CP | 25 (15.8%) | 25 (15.8%) | 5 (11.9%) |

The study subjects with OFCs included 75 male samples (47.5%) and 83 female samples (52.5%). The large majority of OFC participants self-reported their race as White (146, 92.4%), followed by Black (4, 2.5%), multi-racial (3, 1.9%), Asian (2, 1.3%), and American Indian or Alaska Native (3, 1.9%); 7.6% reported Hispanic or Latino ancestry (n=26). Although OFC study subjects ranged in age from 0.01 years (newborn) to 66.5 years; the majority of these subjects were minors (n=122, 77%) whose ages were strongly skewed young as the median age was 4.94 years. Of the 122 minor subjects, 94% (n=115) were under age 12, three-quarters were less than 6 years old (n=90, 74%), more than half were under 4 years (n=69, 57%), and about one-third were age 2 years and younger (n=42, 34%) (Figure 3).

**Figure 3.**
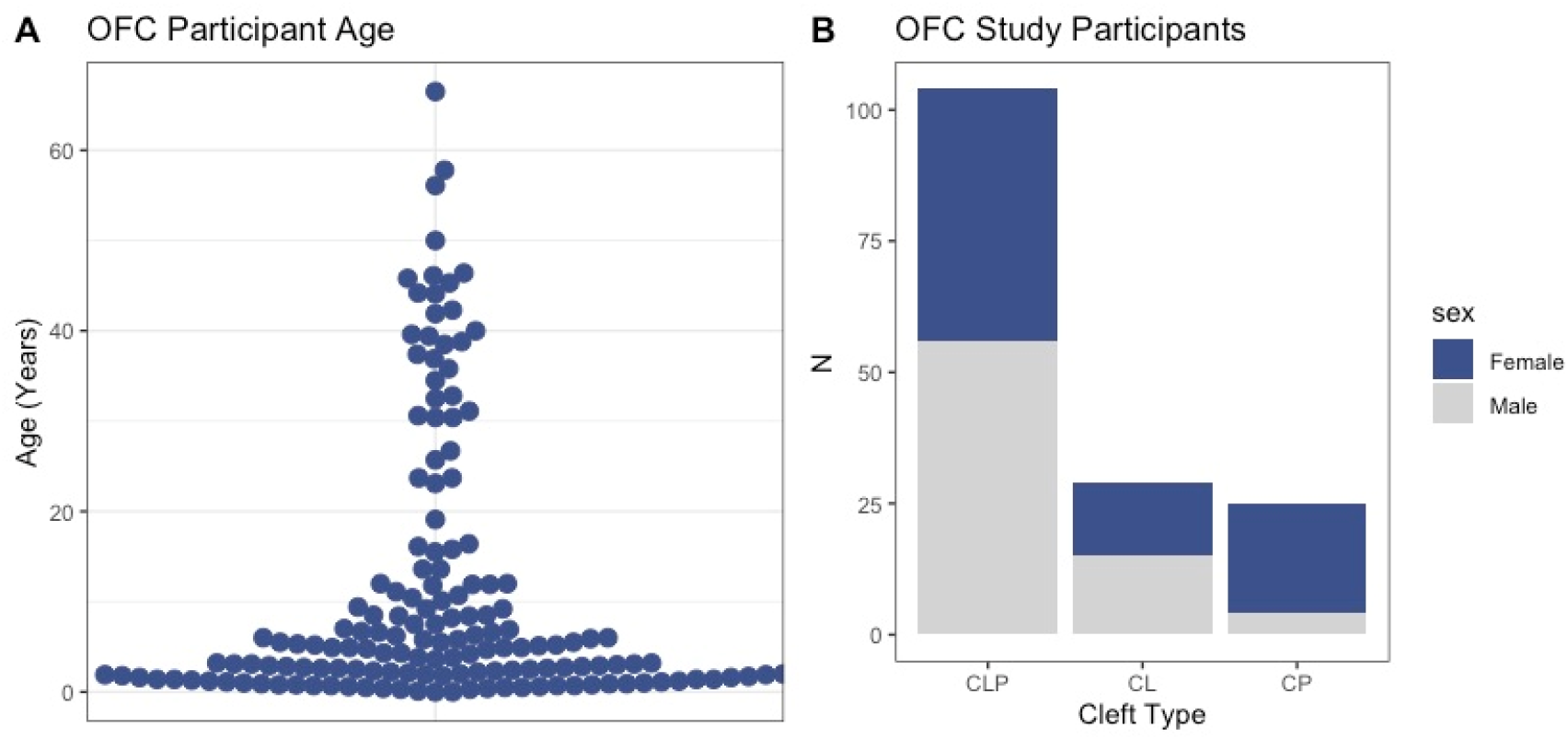
Characteristics of study participants with OFCs. (A) Age of participants with OFCs in years. (B) Sex and cleft types of participants with OFCs.

### OFC Characteristics

There were 262 study subjects with an OFC enrolled in the study, of which 158 study subjects from 130 families returned samples. OFC types were categorized as: cleft lip only, cleft lip and palate, cleft palate only, and unknown. Of the 158 returned OFC samples, cleft types reported were: 18.4% cleft lip only (n=29), 65.8% cleft lip and palate (n=104), and 15.8% cleft palate only (n=25) (Figure 3). The male-to-female ratio for cleft palate only was .21, and 1.23 for clefts involving the lip. Of the 133 total participating families who provided samples, 46 (35%) had a family history of OFC with at least one other known family member affected, and 19 families (14%) reported 2 or more additional OCF-affected family members in addition to the proband. Syndromic clefts were reported for 15 families (11.4%), 10 of which were Van der Woude Syndrome.

### Enrollee Characteristics

The majority of study subjects were minors who did not enroll themselves, but were enrolled in the study by a parent/guardian who self-referred to the study. The characteristics of those who initiated enrollment (“enrollees”) is based on the adult who provided consent-either the affected adult study subject or the parent/guardian of the affected minor study subject. After removing duplicates from families with multiple affected members (multiple children or both parent and child affected) there were 240 unique “enrollees” in the study. The majority of enrollees (94.2%) were female - 226 females compared to 14 males. Age data was available for 181 enrollees, who had an average age of 35.4 years (SD 8.0, range 19.1-72.2 years). 90% of enrollees were less than 45 years old.

### Reasons for participating and participant experience

A feedback survey about study participant experiences was sent to the enrollees of the 133 families who had completed both the online surveys and sample collection, and was completed by 78 (59%). The vast majority of survey respondents found participation in the ECP study to be “very easy” (n=56, 72.7%) or “easy” (n=18, 23.4%), and 100% of survey respondents indicated that they felt they were provided with adequate information about how to participate in the study.

Participants were asked to select their primary reason for participating in the research study. The most common response was “to contribute to scientific knowledge about clefts” (n=44, 56.4%), followed by “to help other/future families” (n=25, 32.1%). Approximately 10% of respondents indicated that their primary motivation was “to learn more about clefts for their family” (n=8, 10.3%), and one participant selected “to receive gift card compensation” as their primary reason for participating. Less than half of respondents (34/78, 43.6%) had previous experience with genetic testing but prior experience with genetic testing either had no influence on the decision to participate in this study (n=19, 55.9%). Interestingly, the one respondent who rated their prior genetic testing experience as “very negative” also denoted that their experience made them “more likely to participate” in this research study.

The feedback survey also asked about potential concerns related to study participation, including logistical concerns (sample collection/return, time commitment), privacy concerns (sharing personal information, DNA sample), personal reservations (religious or ethical concerns) and other concerns.

Most respondents did not have any concerns related to participation (n=69, 88.5%). Concerns around privacy were reported by 6% of participants (n=5), logistical concerns were an issue for approximately 4% (n=3), and one participant indicated personal reservations about participation.

Prior to participating in this research study, approximately 40% of responding participants were moderately or extremely familiar with the research participation process. Approximately 30% of respondents were “not at all familiar” with the research participation process (n=23, 29.5%). 41% of respondents (n=32) had never participated in a research study before.

## Discussion

The goal of this study was to evaluate the feasibility of using social media (SM) to recruit and enroll families in genetic research studies for OFCs. Facebook support groups proved to be an excellent source of participant recruitment, resulting in a large volume of responses from interested families. The support group administrators who responded to our messages were overall interested in helping to promote our study and share research opportunities with their group, with some of them even participating themselves. The endorsement of the study by a leader of the support group likely increased engagement and lent legitimacy to the study for group members who may have previously been unfamiliar with genetic research and/or Emory University. All advertisement for the study was unpaid; as a result, the only cost incurred by the study team was for study staff and supplies. Recruitment using this method was very cost-effective compared to paid advertisement methods or by in-clinic methods, as recruitment was completed with minimal personnel.

The use of an online screening survey significantly streamlined the screening process, automatically creating a record of each interested participant with contact information that could be easily reviewed and connected to a study ID and participant record. This passive form of recruitment in which interested participants completed the survey was found to be very time-efficient, as less time was spent with each participant in the initial screening process to determine eligibility compared to screening individually over phone, email, or in person in a clinical setting. We found the REDCap database to be a critical tool with an ease and flexibility of survey design and automation of the survey distribution and reminder process. Centralization of data in this repository including study communications, study compensation records, and sample logging facilitated comprehensive data management and further automated many administrative processes. REDCap features including audit trails for tracking data manipulation, automated export features to download data to statistical packages, as well as data integration from external sources greatly assisted efficiency of data management.^22,23^ Overall, this recruitment method was a highly effective method to obtain a large number of participants with low overhead costs and time commitment.

Remote recruitment facilitated engagement of a geographically diverse study population. Moreover, we were able to reach and recruit families residing outside close proximity to a major research center, who may have otherwise not had the opportunity to participate in genetic research on OFCs. This was also reflected in the feedback survey where 41% of participants were new to research. Although we were able to recruit a geographically diverse sample cohort from 38 of 50 states, a disproportionate number of participants were located in Georgia and the southeast, which may in part be due to heightened name recognition of Emory University, which is located in the state of Georgia.

Diversity of study participants comes in many forms. Although we were able to recruit a geographically diverse sample, it was overwhelmingly white. Although we did not assess socio-economic status, it is likely that our study is depleted for individuals from lower socio-economic groups. While accommodations could be made to complete study surveys via phone, initial study advertisement was completed entirely online. As a result, few, if any, participants were recruited who did not have access to the Internet, either via mobile device or computer. Furthermore, our study had limited participation by individuals reporting Hispanic ethnicity which is a direct consequence of study materials being only available in English. The lack of racial and ethnic diversity in our study population limits representativeness and generalizability of study results generated from this study population, and points to the persistence of challenges in recruiting adequately diverse study populations, even when recruiting online.

For congenital anomalies, SM support group membership is likely to skew younger as parents would be likely to reach out for support and information during infancy and early childhood, before and/or during their child’s procedure(s), and may be most actively engaged in the support groups during that time. This can present challenges for sample collection and the busy time in the care of an infant/toddler and the treatment of their OFC may contribute to lower return rates. We also found “enrollees” for our study to be overwhelmingly white and female, a problem previously seen in systematic reviews of online study recruitment.^17,21^ However, there was a similar proportion of males and females among OFC individuals, mostly minors, indicating that although mothers are more likely to seek out research participation than fathers, their likelihood of enrolling their child in research does not differ based on their child’s sex. The percentages of each type of OFC in our study participants resembled population frequencies ^2^, indicating that there was no major bias toward one type of OFC or another among SM support groups or in individuals seeking research opportunities.

A major disadvantage of remote recruitment is the high probability of losing participants to follow-up and unreturned sample kits. Each unreturned sample kit increases the cost per participant so low return rates can significantly increase study costs. Because a majority of our minor participants were under the age of 5, obtaining saliva samples can be challenging and require swabs and sponges. Responding to participant feedback and making changes to the kits resulted in a three-fold increase in the number of sample kits returned within 3 weeks from families with minor participants under 2 years of age. Although the self-paced online survey portion of the study proved to be an effective way to involve international participants in the study, there were a number of logistical and cost limitations involved in shipping and receiving samples internationally, compared to domestic samples. Future studies seeking to recruit internationally should consider the significant additional cost and effort involved in ascertaining samples.

Overall, unpaid study advertisement on SM sites was successful in gathering a sizable cohort of geographically diverse participants in a relatively short period of time. Individuals who elected to enroll in the study, for themselves or an affected minor, were overly white and female, however, the affected study participants exhibited the full range of over OFC phenotypes. In the future, study materials may be provided in more than one language, potentially enabling recruitment of more diverse populations. Advertising was free and online recruitment took less time than consenting patients in-clinic, leading to lower personnel costs. The ECP found social media recruitment eminently feasible and an effective component of research study design in the digital age.

## Data Availability

All relevant data is presented in the manuscript.

## Acknowledgements

We sincerely thank all of the families participating in the Emory Cleft Project and our clinical colleagues for referring families to the study. This study was supported by funding from the National Institutes of Health R00-DE025060 and R01-DE028342.

## References

1. Beaty TH, Marazita ML, Leslie EJ. Genetic factors influencing risk to orofacial clefts: today’s challenges and tomorrow’s opportunities. F1000Res. 2016;5:2800.

2. Dixon MJ, Marazita ML, Beaty TH, Murray JC. Cleft lip and palate: understanding genetic and environmental influences. Nat Rev Genet. 2011;12(3):167–178.

3. Hlongwa P, Rispel LC. “People look and ask lots of questions”: caregivers’ perceptions of healthcare provision and support for children born with cleft lip and palate. BMC Public Health. 2018;18(1):506.

4. Cinar S, Boztepe H, Prof FFO. The Use of Social Media Among Parents of Infants with Cleft Lip and/or Palate. J Pediatr Nurs. 2020;54:e91–e96.

5. Statista. Number of monthly active Facebook users worldwide as of 3rd quarter 2020. https://www.statista.com/statistics/264810/number-of-monthly-active-facebook-users-worldwide/. xUpdated November 24, 2020. Accessed December 7, 2020.

6. Sinclair M, McCullough JE, Elliott D, et al. Exploring Research Priorities of Parents Who Have Children With Down Syndrome, Cleft Lip With or Without Cleft Palate, Congenital Heart Defects, or Spina Bifida Using ConnectEpeople: A Social Media Coproduction Research Study. J Med Internet Res. 2019;21(11):e15847.

7. Hausmann JS, Touloumtzis C, White MT, Colbert JA, Gooding HC. Adolescent and Young Adult Use of Social Media for Health and Its Implications. J Adolesc Health. 2017;60(6):714–719.

8. Iliffe LL, Thompson AR. Investigating the beneficial experiences of online peer support for those affected by alopecia: an interpretative phenomenological analysis using online interviews. Br J Dermatol. 2019;181(5):992–998.

9. Khouri JS, McCheyne MJ, Morrison CS. #Cleft: The use of Social Media Amongst Parents of Infants with Clefts. Cleft Palate Craniofac J. 2018;55(7):974–976.

10. Stock NM, Martindale A, Cunniffe C, Research VFRTatCfA. #CleftProud: A Content Analysis and Online Survey of 2 Cleft Lip and Palate Facebook Groups. Cleft Palate Craniofac J. 2018;55(10):1339–1349.

11. Kulanthaivel A, Fogel R, Jones J, Lammert C. Digital Cohorts Within the Social Mediome: An Approach to Circumvent Conventional Research Challenges? Clin Gastroenterol Hepatol. 2017;15(5):614–618.

12. Schumacher KR, Stringer KA, Donohue JE, et al. Social media methods for studying rare diseases. Pediatrics. 2014;133(5):e1345–1353.

13. Comerford M, Fogel R, Bailey JR, Chilukuri P, Chalasani N, Lammert CS. Leveraging Social Networking Sites for an Autoimmune Hepatitis Genetic Repository: Pilot Study to Evaluate Feasibility. J Med Internet Res. 2018;20(1):e14.

14. Lammert C, Comerford M, Love J, Bailey JR. Investigation Gone Viral: Application of the Social Mediasphere in Research. Gastroenterology. 2015;149(4):839–843.

15. Burton-Chase AM, Parker WM, Hennig K, Sisson F, Bruzzone LL. The Use of Social Media to Recruit Participants With Rare Conditions: Lynch Syndrome as an Example. JMIR Res Protoc. 2017;6(1):e12.

16. Timberlake AT, Wu RT, Cabrejo R, Gabrick K, Persing JA. Harnessing Social Media to Advance Research in Plastic Surgery. Plast Reconstr Surg. 2018;142(4):1094–1100.

17. Whitaker C, Stevelink S, Fear N. The Use of Facebook in Recruiting Participants for Health Research Purposes: A Systematic Review. J Med Internet Res. 2017;19(8):e290.

18. Tse RW, Oh E, Gruss JS, Hopper RA, Birgfeld CB. Crowdsourcing as a Novel Method to Evaluate Aesthetic Outcomes of Treatment for Unilateral Cleft Lip. Plast Reconstr Surg. 2016;138(4):864–874.

19. Wu C, Scott Hultman C, Diegidio P, et al. What Do Our Patients Truly Want? Conjoint Analysis of an Aesthetic Plastic Surgery Practice Using Internet Crowdsourcing. Aesthet Surg J. 2017;37(1):105–118.

20. Frandsen M, Thow M, Ferguson SG. The Effectiveness Of Social Media (Facebook) Compared With More Traditional Advertising Methods for Recruiting Eligible Participants To Health Research Studies: A Randomized, Controlled Clinical Trial. JMIR Res Protoc. 2016;5(3):e161.

21. Thornton L, Batterham PJ, Fassnacht DB, Kay-Lambkin F, Calear AL, Hunt S. Recruiting for health, medical or psychosocial research using Facebook: Systematic review. Internet Interv. 2016;4:72–81.

22. Harris PA, Taylor R, Minor BL, et al. The REDCap consortium: Building an international community of software platform partners. J Biomed Inform. 2019;95:103208.

23. Harris PA, Taylor R, Thielke R, Payne J, Gonzalez N, Conde JG. Research electronic data capture (REDCap)--a metadata-driven methodology and workflow process for providing translational research informatics support. J Biomed Inform. 2009;42(2):377–381.

